# Parameter Estimation of COVID-19 Pandemic Model with Self Protection Behavior Changes

**DOI:** 10.1101/2020.08.24.20180695

**Authors:** Kassahun Getnet Mekonen, Tatek Getachew H/Michael, Shiferaw Feyissa

## Abstract

A mathematical model for the transmission dynamics of Coronavirus diseases (COVID-19) is proposed by incorporating self-protection behavior changes in the population. The disease-free equilibrium point is computed and its stability analysis is studied. The basic reproduction number(*R*_0_) of the model is computed and the disease-free equilibrium point is locally and globally stable for *R*_0_ < 1 and unstable for *R*_0_ *>* 1. Based on the available data the unknown model parameters are estimated using a combination of least square and Bayesian estimation methods for different countries. Using forward sensitivity index the model parameters is carried out to determine and identify the key factors for the spread of disease dynamics. From country to country the sensitive parameters for the spread of the virus varies. It is found out that the reproduction number depends mostly on the infection rates, the threshold value of the force of infection for a population, the recovery rates, and the virus decay rate in the environment. It is also demonstrated that control of the effective transmission rate (recommended human behavioral change towards self-protective measures) is essential to stop the spreading of the virus. Numerical simulations also show that the virus’s transmission dynamics depend mostly on those sensitive parameters.

## 1 Introduction

The outbreak of coronavirus was first informed to the World Health Organization (WHO) as pneumonia of unknown cause on December 31, 2019 in Wuhan City, Hubei Province, China. As of 10 January 2020, the virus causing the outbreak was further determined by gene sequencing to be the new novel coronavirus, the same category as the Middle Eastern Respiratory Syndrome virus (MERS-CoV) and the Severe Acute Respiratory Syndrome virus (SARSCoV) [16]. On 30 January 2020, the epidemic of coronavirus disease 2019 (COVID-19) was declared as a public health emergency of international concern, the highest level in the emergency response for infectious diseases [9]. The rapid spread of this virus with consequences on an international scale, COVID19 was declared a pandemic by the WHO on March 11, 2020 [1]. The global report of COVID-19 by WHO indicated that on August 16, 2020, above 21.2 million people were infected with the virus and over 761,779 were died [22]. The outbreak of the disease is still rapidly increasing in South American, North American, Asian and African Countries at an alarming rate.

The novel coronavirus is a respiratory virus that spreads primarily through droplets of saliva or discharge from the nose generated when an infected person coughs or sneezes [1]. Individuals can also be infected from contacting surfaces contaminated with the virus and touching their eyes, nose and mouth. The COVID-19 virus may survive on surfaces for long periods with the right environmental conditions [23]. Understanding the transmission characteristics of the diseases in communities, regions, and countries leads to better approaches to decrease the transmission of these diseases [12].

There is no known curing medicine nor vaccine to combat the COVID-19 pandemic. However, most symptoms can be treated and getting early care from a healthcare provider can make the disease less dangerous. There are several clinical trials that are being conducted to evaluate potential therapeutics for COVID-19 [23]. Standard recommendations by WHO to prevent the spread of COVID-19 include frequent cleaning of hands using soap or alcohol-based sanitizer, covering the nose and mouth with a flexed elbow or disposable tissue when coughing and sneezing, and avoiding close contact with anyone that has a fever and cough [23]. The awareness of individuals for applying these preventive mechanisms vary from region to region and from country to country. In some places, protective measures are employed by volunteer individuals while in some other places, governments impose some kind of rules on the population to use strict physical distancing and wearing face masks at public places [14].

Mathematical models have become important tools in understanding and analyzing the spread and control of infectious diseases, which clarifies variables and parameters to obtain conceptual results such as basic reproduction numbers [24]. Mathematical models and computer simulations are useful for determining sensitivities to change in parameter values, and estimating key parameters from data which can contribute to identifying trends, make general forecasts, and estimate the uncertainty in forecasts [12]. Understanding the virus dynamics and host response is essential in formulating strategies for antiviral treatment, vaccination, and epidemiological control of COVID-19 [7]. The analysis from mathematical models may assist decision makers to estimate the risk and the potential future growth of the disease in the population [14].

Since epidemiological and mathematical models play a fundamental role in the study of the dynamics of such COVID-19, various models have been used to investigate the transmission dynamics of the pandemic [4, 18, 13, 8, 27, 25, 11, 32]. In [25] a generalized SEIR model was developed to analyze this epidemic. Based on the public data of the National Health Commission of China from Jan. 20th to Feb. 9th, 2020, they estimate epidemic parameters and make predictions on the possible ending time for 5 different regions.

Behavior change towards using preventive mechanisms by the population to protect themselves from an infectious disease are assumed to be dependent on the way that the disease is transmitted and its fatality [15]. Individuals who have awareness about the disease and decided to use preventive mechanisms have less susceptibility than those without awareness and demonstrating the usual risky behavior [14, 30]. In this paper, we propose SEIRDM mathematical model for the transmission dynamics of COVID-19 by introducing a behavior change function.

In order to get better predictions and to design and analyse various intervention strategies, one needs to estimate the model parameters from existing epidemiological data. It is most unusual to estimate parameter values from observed data in dynamical systems. But some authors estimate parameters based on the available data to determine the effects of various epidemiological factors on disease transmission and possible control strategy. Least Squares Support Vector Machines for parameter estimation of time invariant and time varying dynamical systems are used in [20]. Biegler and Grossmann [2] employed optimization techniques based on the seasonal data with the SIRS epidemic model in order to estimate the parameters of a generalized incidence rate function. The SIR model parameters were numerically estimated in [5] using the least squares method. In our case, the model parameters are inferred or estimated from observed data for different countries use a modified Bayesian approach combined with least square techniques. Sensitivity analysis of the model parameters is carried out to determine and identify the key factors for the spread of disease dynamics. We use forward sensitivity analysis to identify the most sensitive parameters.

The paper is organized as follows: The mathematical model is formulated and described in Section 2. The qualitative analysis of the model by examining the equilibrium points and its stability analysis is studied in Section 3. Numerical simulations of the model by estimating the parameters are given in Section 4. The sensitivity analysis of the basic reproduction number for the model with respect to the parameters is also discussed in this section. Conclusions and recommendations of the study are given in Section 5.

## 2 Model Formulation

The model subdivides the human population into five disjoint compartments; Susceptible, Exposed, Infected, Recovered, death; and one compartment which is the contaminated material or surface. We consider the following basic assumptions to formulate the model.

1. The transmission dynamics of COVID-19 is similar to the SEIR model. We add the death compartment for which individuals die via the virus, in account with they may also die via the natural death.
2. We consider the contribution of the asymptomatic infectious individuals in the transmission dynamics of disease in the population.
3. The effect of indirect transmission of the disease through virus concentration in the environment due to shedding by infectious is considered.
4. We apply behavior change towards self-protective measures by the population to protect themselves from the virus.

We will then propose a mathematical model and analyze the effect of these factors to investigate in terms of their contribution to prevent the spread of disease.

The model state variables and parameters with their meanings are given in Table 1 and Table 2 respectively.

**Table 1:** The Subdivided Compartments in our Model

| Symbol | Biological Description of the State Variables |
| --- | --- |
| $S$ | Susceptible individuals |
| $E$ | Latently infected individuals, who have no symptoms of COVID-19 virus disease and are infectious |
| $I$ | Infected individuals, who have active disease and are infectious |
| $R$ | Recovered individuals |
| $D$ | Individuals who died only with COVID-19, but not with other disease and natural deaths |
| $M$ | COVID-19 contaminated materials or surfaces in the environmnet |

**Table 2:** Description of the model parameters

| Parameter | Biological Description of the parameter |
| --- | --- |
| $\Lambda$ | Rate of recruitment to the susceptible individuals |
| $\alpha$ | Rate of dissemination of information about the disease in the population |
| $\beta_1$ | Rate of disease transmission directly from humans |
| $\beta_2$ | Rate of disease transmission from the environment |
| $\lambda$ | Force of infection ( It is the probability of acquiring infection from an infected individual) |
| $\lambda_0$ | Threshold value of the force of infection for a population to start reacting swiftly |
| $K$ | the pathogen concentration in the environment that yields 50% of chance for a susceptible individual to catch the viral infection from the environment |
| $\nu$ | Modification Parameter |
| $e$ | Behavior change function |
| $\theta$ | Rate of recovery of the individuals from Exposed class |
| $\eta$ | The average effectiveness of existing self-preventive measures |
| $\gamma$ | Rate of recovery of the individuals from virus in the Infected class |
| $\delta$ | Death rate due to the virus |
| $\mu$ | Natural death rate of the individuals |
| $\psi$ | Decay rate of the virus from the environment |
| $\epsilon$ | Shedding rate of the virus from the exposed class to the environment |
| $\xi$ | Shedding rate of the virus from the infected class to the environment |

The total population at time t, denoted by *N*(*t*), is given by

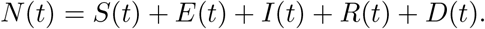

The flow diagram of the model is illustrated in Figure 1.

Based on our assumptions and the flow diagram, it results in systems of the following non-linear differential equations:

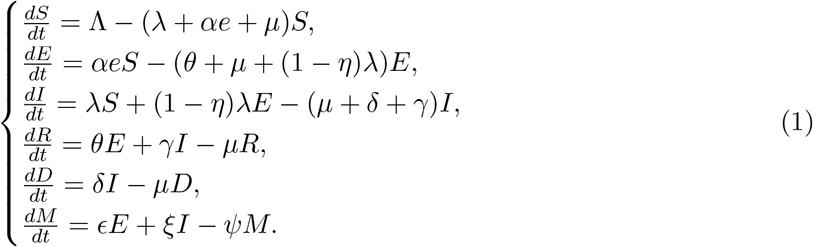

**Figure 1:**
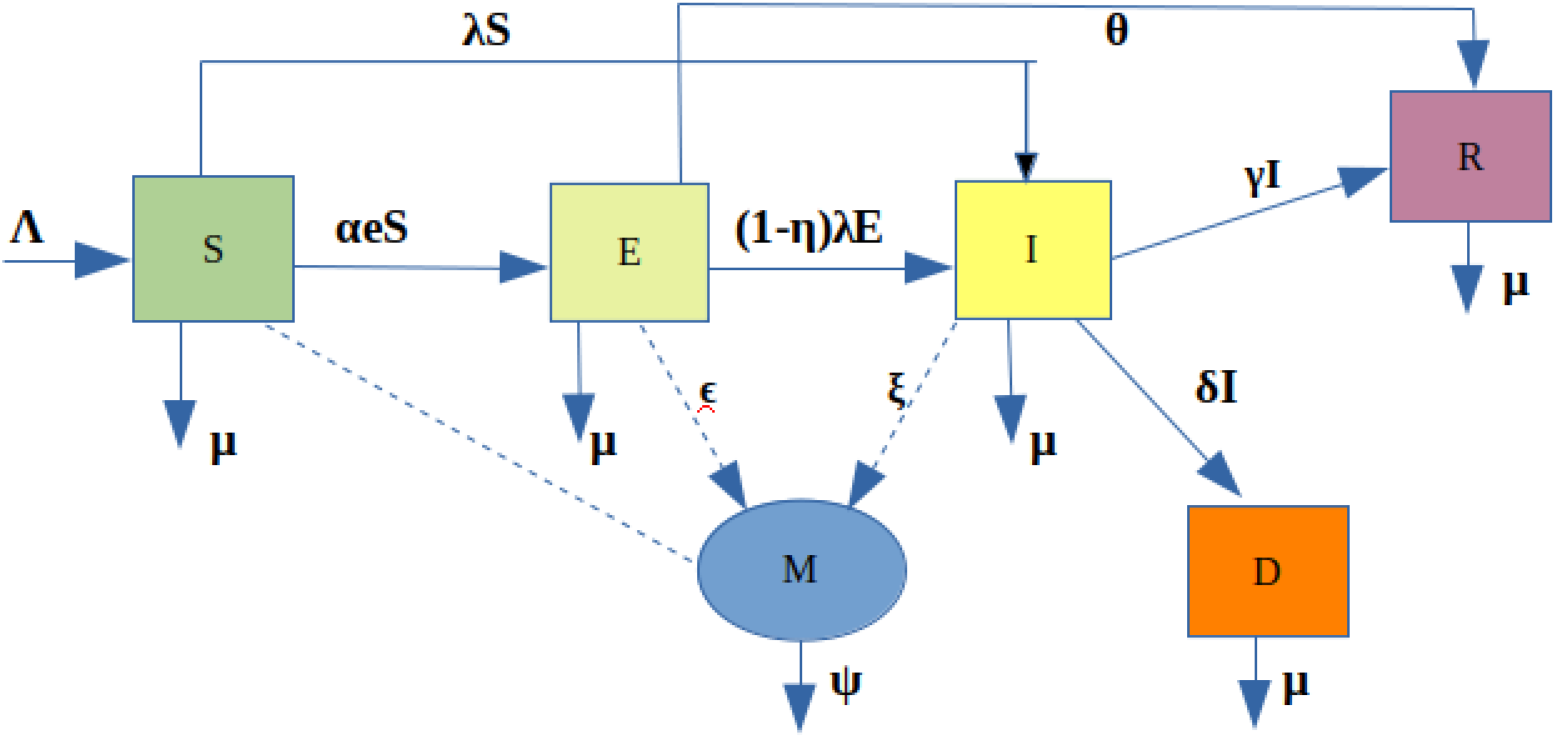
The flow Diagram for the Model of COVID-19 Pandemic

with non negative initial conditions *S*(0) *>* 0, *E*(0) ≥ 0, *I*(0) ≥ 0, *R*(0) ≥ 0, *D*(0) ≥ 0 and *M*(0) ≥ 0.

### 2.1 Model Description

The susceptible population is increased by the recruitment of individuals into the population, at a rate *Λ*. All human individuals suffer from natural death, at a constant rate *µ*. Susceptible individuals acquire COVID-19 infection from individuals with exposed classes at a rate *αe* and the infected class at a rate *λ*. The behavior change function *e* and the force of infection *λ* are given by

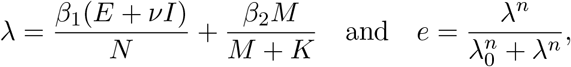

where *β*_1_ is the rate of virus transmission directly from humans, *β*_2_ is the rate of virus transmission from the environment, *K* is the pathogen concentration in the environment that yields 50% of chance for a susceptible individual to catch the viral infection from the environment, *λ*_0_ is the value of the force of infection corresponding to the threshold infectivity in which individuals start reacting swiftly (that means, the point at which the behavior change function changes its concavity) and *n* is a hill coefficient that portrays the rate of reaction by the population. The modification parameter *ν* ≥ 1 accounts for the relative infectiousness of individuals with COVID19 symptoms, in comparison to those infected with the virus and with no symptoms. Individuals with virus symptoms are more infectious than that of without symptom because they have a higher viral load and there is a positive correlation between viral load and infectiousness [29].

At the beginning of an outbreak, individuals understand very little about the virus; there could be no reaction and this can be related to the situation at the disease-free equilibrium such that *e*(*λ*)= *e*(0) = 0. However, as the risk of the disease increases, individuals start to think of the type of measures to take in order to avoid all means of contracting the disease. These protection measures, if perfect, account for an increase in the values of *e* to unity. The order *n* of the function *e*(*λ*) is a Hill coefficient that portrays the rate of reaction by the population [15].

The rate of dissemination of information *α* describes the awareness of individuals from the disease. Individuals acquire information through multiple ways, such as TV news, reports on a network, mouth-to-mouth communication, or even education. The information individuals gathered is an essential factor which impacts how individuals react to the disease transmission and individuals make behavior changes to keep themselves from infection based on the information available to them during the pandemic [35].

Individuals leave the exposed class *E* by becoming symptomatic, at a rate (1 − *η*)*λ* with the average effectiveness of existing self-preventive measures *η* ≤ 1, or recovered at rate *θ*. Individuals with the symptoms of COVID-19 disease dies due to the virus-induced death at a rate *δ*. The infected individuals also become recovered at a rate *γ*. We assume that the recovered individuals *R* acquire partial immunity. The released COVID-19 from the exposed and infected individuals through coughing or sneezing landed on materials or surfaces around them and become infected at a rate *ϵ* and *ξ*, respectively [14, 30]. The virus decays from the infected surfaces with the decay rate of *ψ*.

## 3 Analysis of the Model

In this section, we will see the qualitative analysis of the model Equation (1) by examining the equilibrium points and its stability analysis.

### 3.1 Well-Posedness

Let us begin understanding the dynamics of a model by examining the behavior about its steady states. We first show that the model is well posed in a biologically feasible domain, and then proceed with a stability analysis of the steady states of the model.

#### Theorem 3.1

1. *There exists a unique solution to the system of equations (1) in the region* 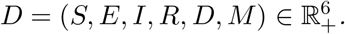
2. *If S*(0) *>* 0*,E*(0) *≥* 0*,I*(0) ≥ 0*,R*(0) *≥* 0*,D*(0) *≥* 0 *and M*(0) ≥ *0, then S(t) >* 0*,E(t) ≥* 0*,I(t)* ≥ 0*,R(t)* ≥ 0*,D(t)* ≥ *0 and M(t)* ≥ 0 *for all t* ≥ 0.
3. *The solution trajectories of the model equations (1) evolve in abounded and positive invariant region*

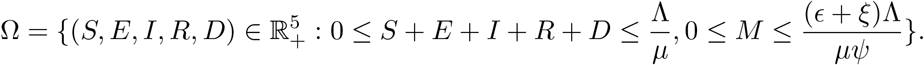

##### Proof

The Well-Posedness of the Model is proved as follow:

1. All the functions on the right hand side of Equation (1) are *C*^1^ on ℝ^6^. Thus, by the Picard–Lindelöf theorem [28], equations (1) has a unique solution.
2. The positivity of model state variables are proved based on proposition A.1 in [31]. Let the model equations (1) be written in the form *x*′= *F* (*x, t*), where *x* =(*S, E, I, R, D, M*), and 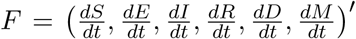. The functions *F* (*x, t*) on the right hand side of equations (1) have the property of *F* (*S, E, I, R, D, M, t*) ≥ 0 whenever *x* ∈ [0, ∞)^n^ *,x_j_* =0*,t* ≥ 0. Here our *x_j_*’s are *x*_1_ = *S, x*_2_ = *E, x*3 = *I, x*4 = *R, x*5 = *D* and *x*6 = *M*. By Theorem 3.1 (1), there exists a unique solution for the model Equations (1). Thus, it follows from the Proposition that *x*(*t*) ∈ [0, ∞)^n^ for all *t* ≥ *t*0 ≥ 0 whenever *x*(*t*0) ≥ 0.
3. The change of total population *N*(*t*)= *S*(*t*)+ *E*(*t*)+*I*(*t*)+ *R*(*t*)+ *D*(*t*) at time t is governed by *N′*(*t*)= *S′*(*t*)+ *E′*(*t*)+ *I′*(*t*)+ *R′*(*t*)+ *D′*(*t*). That is:

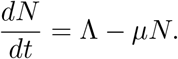 The solution for this linear first order ode is 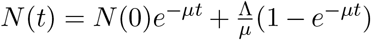. Thus, for the initial data 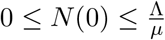, we obtain

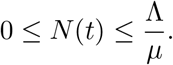 Moreover, for the environmental variable *M*, we have

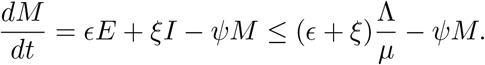 As *E*(*t*) and *I*(*t*) are less than 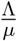. Using the same procedure or by applying the Gronwall inequality, for 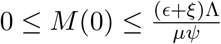, we obtain:

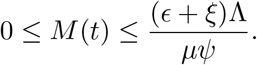 If *x*_0_ is a point in *D*, then the solution of the initial value problem (1), exists for all times *t* ≥ 0 by 3.1 (1). By the result of Theorem 3.1 (2), the solution lies in *D*, for all *t* ≥ 0. Hence the region *Ω* is positive invariant.

Then we will analyze the model quantitative behaviors in the domain *Ω*.

### 3.2 Local Stability of Disease-free Equilibrium (DFE)

The equilibrium solutions of the model are obtained by setting the right-hand side of equation (1) equal to zero:

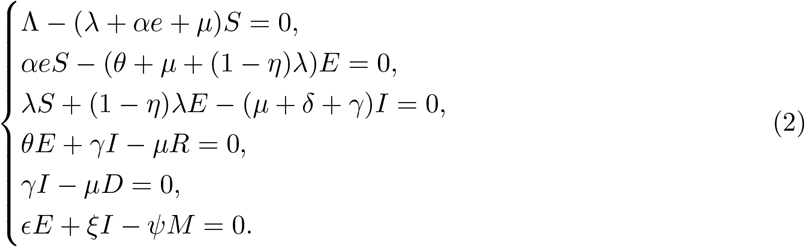

The disease-free equilibrium point of our model is obtained by setting the disease state variables *E* = 0 and *I* = 0. If *E* = 0 and *I* = 0, then *R* = 0 and *D* = 0. It is then denoted and given by:

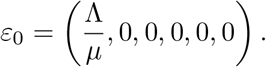

Now let us calculate basic reproduction number denoted by *R*_0_, and defined as the average number of secondary infections produced by a single infected individual in a totally susceptible population. Using the next generation matrix method [33], the basic reproduction number is calculated as follows. From the model equation (1), using the notation *X* =(*E, I, M*), we have the vector functions:

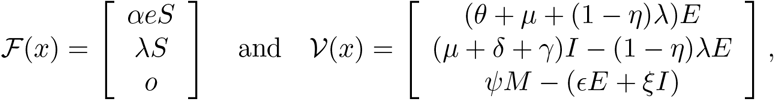

representing the appearance of new infections, and the transfer of individuals in to and out of the infected compartments, respectively. The Jacobian matrices of 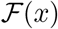 and 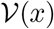 are, respectively

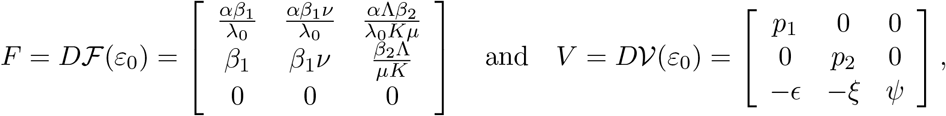

where *p*_1_ = *μ* + *θ*, *p*_2_ = *μ + δ + γ* and the entries of *F* and *V* are obtained by 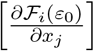 and 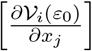 respectively. It is easy to calculate the inverse of *V* and given by

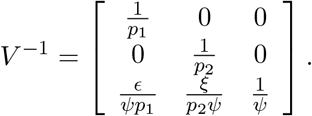

The next-generation matrix *FV*^−1^ is

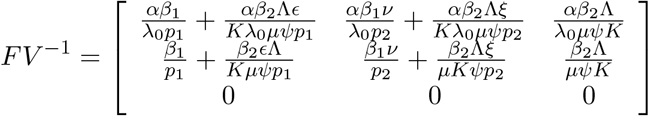

and its eigen values are *λ*_1_ = 0, *λ*_2_ = 0 and 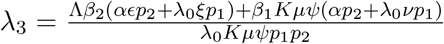. The spectral radius (the largest eigen value) of the next generation matrix is the basic reproduction number of the model. Hence we have the following result.

#### Theorem 3.2

*The basic reproduction number of the model equation (1) is given by*

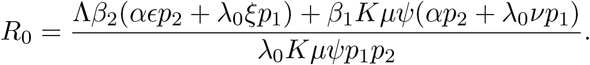

#### Remark 3.3

*In general, if R*_0_ *>* 1*, then, on average, the number of new infections resulting from one infected individual is greater than one. Thus, COVID-19 infections will persist in the populations. If R*_0_ < 1*, then, on average, the number of new infections generated by one infected individual is less than one. This implies that the infections will eventually disappear from the populations. This threshold can as well be used to depict parameters which are most important during the infection*.

The local stability analysis of the equilibrium points are analyzed using linearization. For *n* = 1, the expression *αe* is simplified as 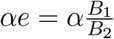, where,

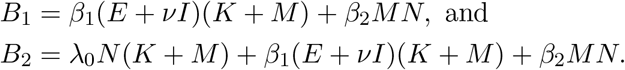

The Jacobean matrix of the model equation (1) is

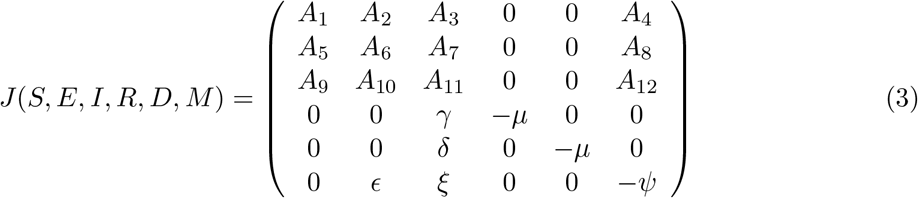

where,

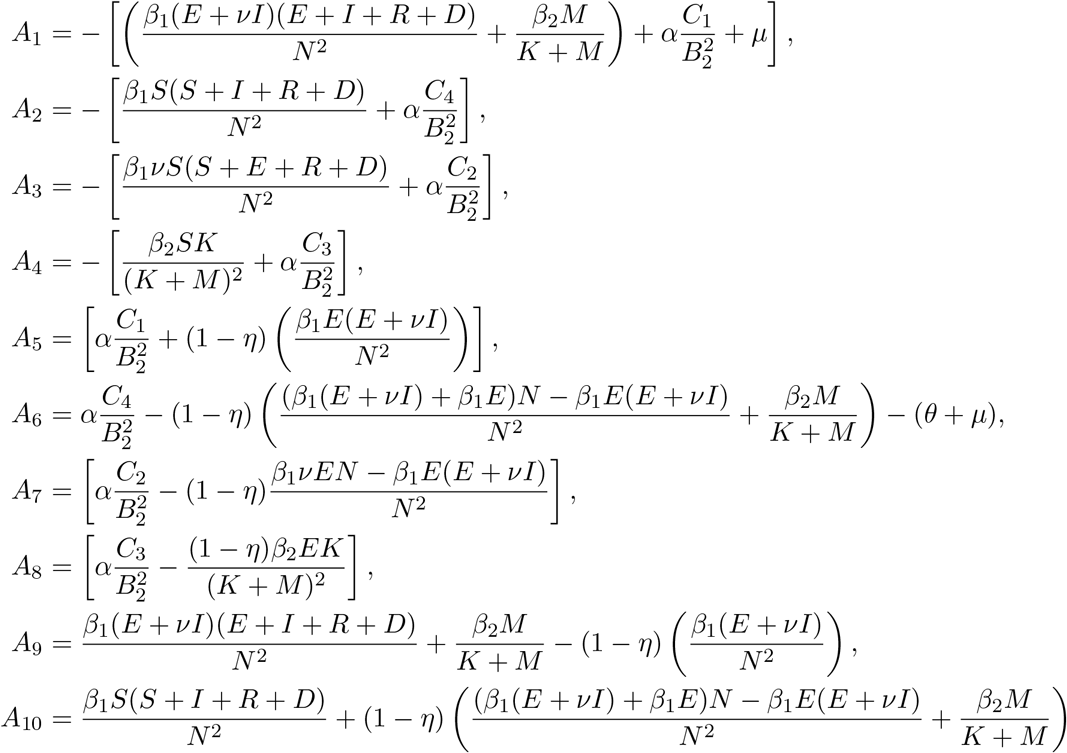

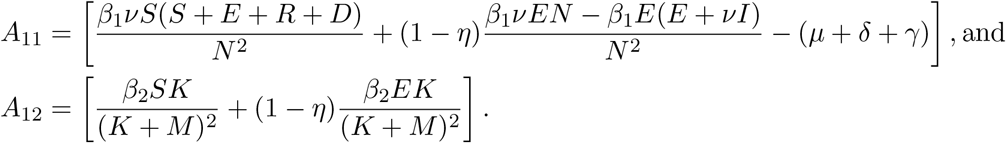

Here

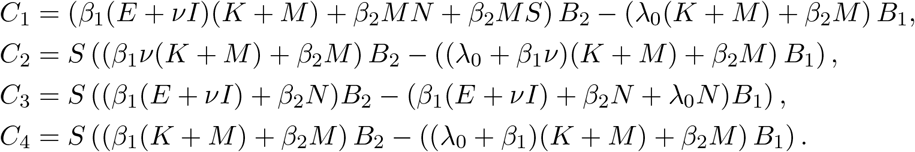

#### Theorem 3.4

*The disease-free equilibrium point ε*_0_ *is locally asymptotically stable if R*_0_ < 1 *and unstable if R*_0_ *>* 1.

##### Proof

The locally asymptotical stablility of *ε*_0_ is obtained use the sign of the eigenvalues of Jacobian matrix at the disease-free equilibrium point *ε*_0_. Substituting the equilibrium point with 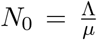 (at the disease-free equilibrium, the total population *N* is equal to the total susceptible *S* population) in the matrix equation (3), we have:

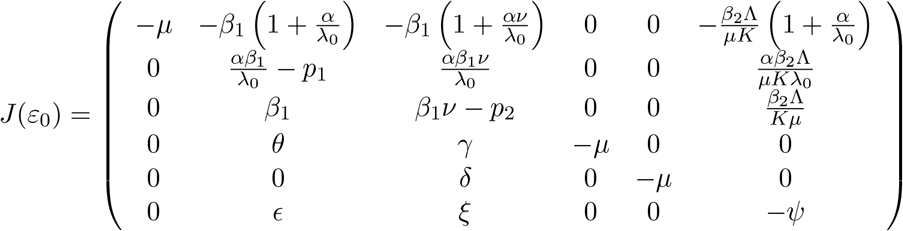

By expanding the characteristic equation |*λI* −*J*(*ε*_0_)| = 0 with the first, fourth and fifth columns, we obtain three eigenvalues *λ*_1_,_2,3_ = −*µ*. The remaining three eigenvalues are obtained from the reduced matrix

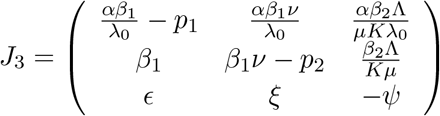

The characteristic equation of *J*_3_ is a third degree polynomial which is given by:

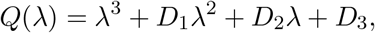

where

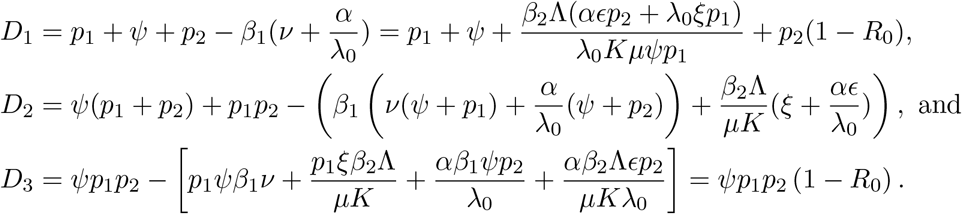

The sign of *D*_1_, *D*_2_ and *D*_3_ are positive if *R*_0_ < 1. It is also true that *D*_1_*D*_2_ *>D*_3_. Using Routh-Hurwitz stability criterion the disease-free equilibrium point *ε*_0_ is stable if *R*_0_ < 1 and unstable if *R*_0_ *>* 1.

### 3.3 Global Stability of Disease-free Equilibrium

Let us rewrite our model system (1) as

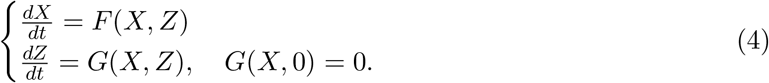

where *X* =(*S, R, D*) and *Z* =(*E,I,M*), with the components of 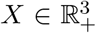 denoting the number of uninfected individuals and 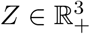 denoting the number of infected ones [6]. The disease-free equilibrium is denoted now as

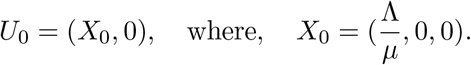

The conditions (*H*_1_) and (*H*_2_) below must be met to guarantee global asymptotically stability: (*H*_1_) For 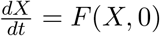, *U*_0_ is globally asymptotically stable;

(*H*_2_) *G*(*X, Z*)= *AZ* − *Ĝ*(*X, Z*)*,Ĝ*(*X, Z*) ≥ 0 for (*X, Z*) ∈ *Ω*, where *A* = *D*_Z_ *G*(*U*_0_, 0) is a Metzler matrix (the off diagonal elements of *A* are non-negative) and *Ω* is the region where the model makes biological sense.

#### Theorem 3.5

*The disease-free equilibrium point U*_0_ =(*x*_0_, 0) *is a globally asymptotically stable equilibrium of (1) if R*_0_ < 1 *and the assumptions* (*H*_1_) *and* (*H*_2_) *are satisfied*.

##### Proof

We have

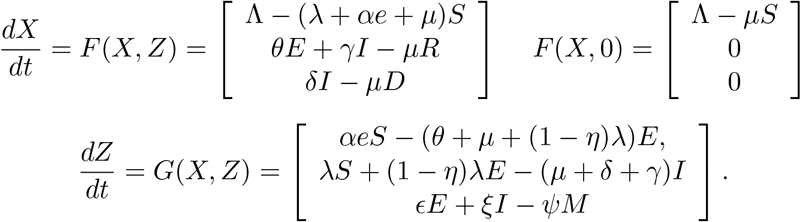

Therefore,

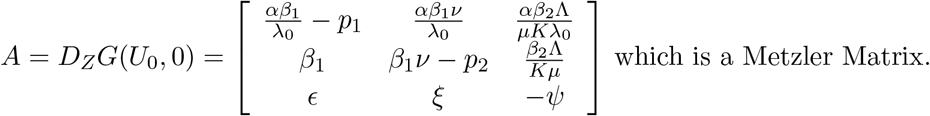

Here *Ĝ*(*X, Z*)= *AZ* − *G*(*X, Z*), and so,

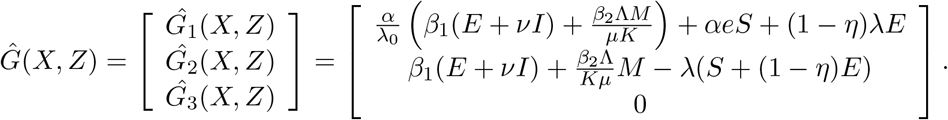

Since 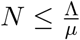 and (1 − *η*) ≤ 1,

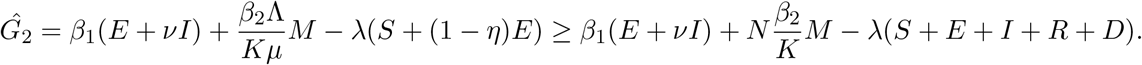

Taking *N* as a common factor, implying

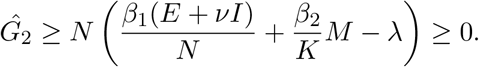

It follows that *Ĝ*_1_(*X, Z*) ≥ 0*,Ĝ*_2_(*X, Z*) ≥ 0 and *Ĝ*_3_(*X, Z*) = 0. Thus, *Ĝ* ≥ 0. Conditions (*H*_1_) and (*H*_2_) are satisfied, and we conclude that *U*_0_ is globally asymptotically stable for *R*_0_ < 1. □

## 4 Parameter Estimation and Numerical Simulations

In this section, we discuss and estimate the parameter choices and the numerical solutions of the model equations (1). We outline the initial conditions and fit the existing WHO data with the model for that choice of parameters.

### 4.1 Parameter Estimation

The systems of model equations (1) can be expressed as a dynamical system of the form:

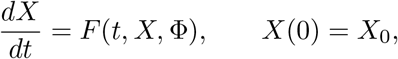

where *t* is the independent variable (time), *X* is the state vector of the system, 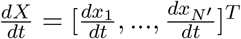, *X* =[*x*_1_*,…,xN*′], *F* =[*f*1*, …, fN*′] and *N′* is the number of compartments in the population. Φ=[ϕ_1_*,…*,ϕ*_p_*] are *p* unknown parameters of the system and *x*_0_ are the initial values [20].

In order to estimate the unknown parameters Φ, the state variable *X*(*t*) is observed at *L* time instants *t*_1_*,…,t_T_*, so that we have

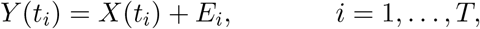

where *Y* (*t_i_*) is the observed values of the state variables at time instant *t_i_* and 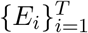 are the difference between the observed value *y_i_* and the corresponding fitted value *x_i_* i.e., *E_i_* = *y_i_* − *x_i_*. The objective is to determine appropriate parameter values so that the sum of squared errors between the outputs of the estimated model (*X*(*t*)) and the measured data (*Y* (*t*)) should be minimized.

We wish to find the vector of least-square estimators, Φ, that minimizes

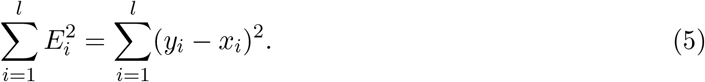

To find the values of parameters Φ that minimizes Equation (5), various methods have been used for handling this problem. The first technique is to differentiate 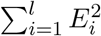 with respect to each Φ and set the results equal to zero to obtain a system of equations that can be solved simultaneously for the Φ’s [21].

To estimate the model parameters we use two step approach

a. The first approach requires solving the ordinary differential equation (1). As using analytic methods are difficult, we the numerical techniques to solve ode’s like runge-kutta methods.
b. The second approach is finding the optimization algorithm to update the parameters based on equation (5)

The process of updating the parameters continues until no significant improvement(convergence) in the objective function is observed. We use a type of Bayesian technique to estimate the unknown parameters and to solve the optimization algorithm.

In this parameter estimation procedure, we modified and combined the Bayesian estimation technique with methods of least square technique. The acceptance and rejection procedure in Metropolis Hasting (MH) algorithm is replaced by comparing the minimum of the sum squared errors between the proposed parameter and the previously assigned parameter. Φ = [ϕ_1_*, …*, ϕ*_p_*] is a vector of parameters, where, *p i*s the number of parameters to be estimated. In our case the number of parameters is *p* = 15. We take one parameter at a time and consider the other parameters held constant in the objective function,

i.e

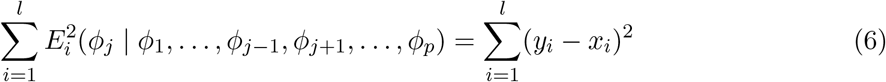

This is a kind of Gibbs Sampling technique for parameter estimation [10]. By combining these parameter estimation techniques we estimate all vector of parameters until convergence.

First, we have to initialize the parameters in their parameter space and propose the next parameter value by sampling from the proposal density. The proposal density we assign 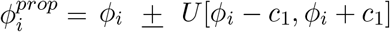, where *c*_1_ is tuning value which is a small number and help us to move the parameter *ϕ_i_* up and down through the parameter estimation process. A rough outline of the algorithm is given in Algorithm (1).

#### Algorithm 1

MH algorithm with Least square Input: *B* (number of iteration), *ϕ*^0^ (initial value for parameters), *Y* (Observed data)

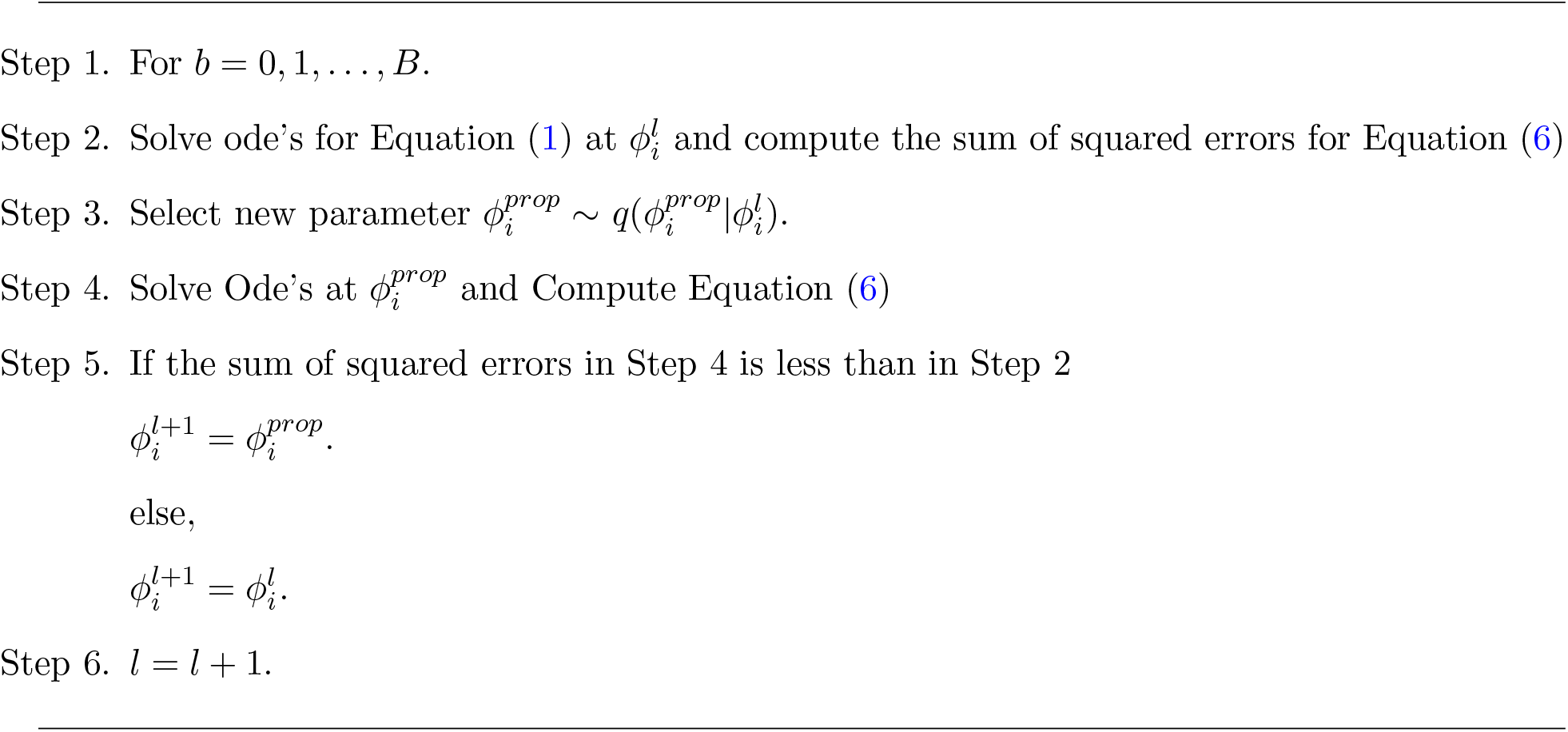

Convergence analysis of the parameter estimation is assessed by line plots of separate parameters.

### 4.2 Numerical Results and Discussion

The parameters must be estimated and assigned a value in order to make the model operable. In this paper, we use total active cases, totally recovered and total death data extracted from WHO situation reports 1 − 192, and worldometer [34], with the daily data from January 21, 2020 to August 16, 2020. We use this data in parameter estimation for countries such as China, Italy, Brazil, South Africa, and Ethiopia. We denote the observed data ***Y*** =[*I, R, D*] with having different length of time *T fo*r those countries. The goal is to find the value of the parameters which minimize the squared errors between the model predictions and the observed data. We also use different initial states of the dynamics for each of the countries. We take initial values for Infected(*I*_0_), Recovered(*R*_0_) and Death(*D*_0_) cases reasonably the same as the observed data *Y*_0_, and we make the assumption that about 80% of the disease is asymptomatic which helps us to outline the initial value for Exposed(*E*_0_). By an initial guess of the parameters *ϕ*^0^ we use *B* = 10*, 00*0 number of iterations for estimation in the MH algorithm.

Based on the available data and the prediction of the proposed model the error terms are computed at the three state compartments. The parameter update is based on the minimization of sum squared differences between measurements and the model predictions.

The convergence analysis of some estimated parameters is given in Figure (2) and our parameter estimation algorithm seems to converge at 2,000 iterations. We take the values at the final iteration as the estimated parameter value for the proposed model. The estimated values of parameters for the countries China, Italy, Ethiopia, South Africa, and Brazil are given in Table (3).

**Figure 2:**
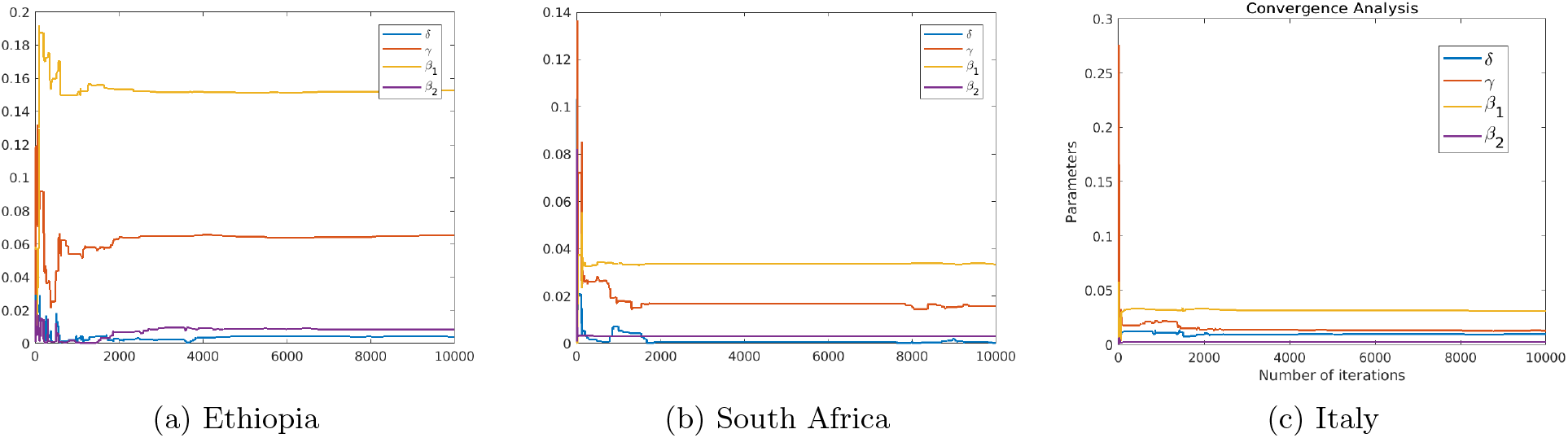
Convergence analysis of a sample of parameters for the COVID-19 induced death rate *δ*, recovery rate *γ* and the contact rates *β*_1_, *β*_2_.

**Table 3:** Estimated Parameter Values at the End of 10,000 Iterations

| Parameter | Estimated Value |  |  |  |  |
| --- | --- | --- | --- | --- | --- |
|  | China | Italy | Ethiopia | Brazil | South Africa |
| $\alpha$ | 0.9865 | 0.8642 | 0.4224 | 0.6594 | 0.2459 |
| $\mu$ | 0.0078 | 0.0122 | 0.0430 | 0.0022 | 0.0233 |
| $\eta$ | 0.9998 | 0.9999 | 0.0986 | 0.0962 | 0.0725 |
| $\delta$ | 0.0039 | 0.0105 | 0.0041 | 0.0043 | 0.0018 |
| $\gamma$ | 0.0326 | 0.0200 | 0.0652 | 0.0461 | 0.0069 |
| $\epsilon$ | 0.0075 | 0.0755 | 0.0021 | 0.0155 | 0.2355 |
| $\xi$ | 0.0323 | 0.0151 | 0.0027 | 0.0081 | 0.0002 |
| $\psi$ | 0.7862 | 0.7224 | 0.6968 | 0.8808 | 0.5788 |
| $\beta_1$ | 0.0801 | 0.0328 | 0.1527 | 0.0116 | 0.0407 |
| $\beta_2$ | 0.0089 | 0.0023 | 0.0083 | 0.0003 | 0.0020 |
| $\lambda_0$ | 0.1062 | 0.071756 | 0.2125 | 0.0541 | 0.0437 |
| $K$ | 437.9314 | 674.8087 | 576.9424 | 842.5115 | 874.1602 |
| $\Lambda$ | 105.9383 | 1195.8673 | 1907.7356 | 712.7638 | 1128.3940 |
| $\theta$ | 0.0064 | 0.0105 | 0.0011 | 0.1892 | 0.1939 |
| $\nu$ | 1.0023 | 1.0015 | 1.0096 | 4.3520 | 1.8138 |

From the estimated parameter values in Table (3) the natural death rate *µ* is relatively higher in Ethiopia and South Africa while the induced death rate due to COVID-19 *δ* is relatively higher in Italy.

Figures (3) shows the fitted model with the observed data for South Africa. The estimated parameters for the infected compartment is well fitted and approximated compared with the observed data. It is also shown that the recovered and death compartment has relatively small under predictions.

**Figure 3:**
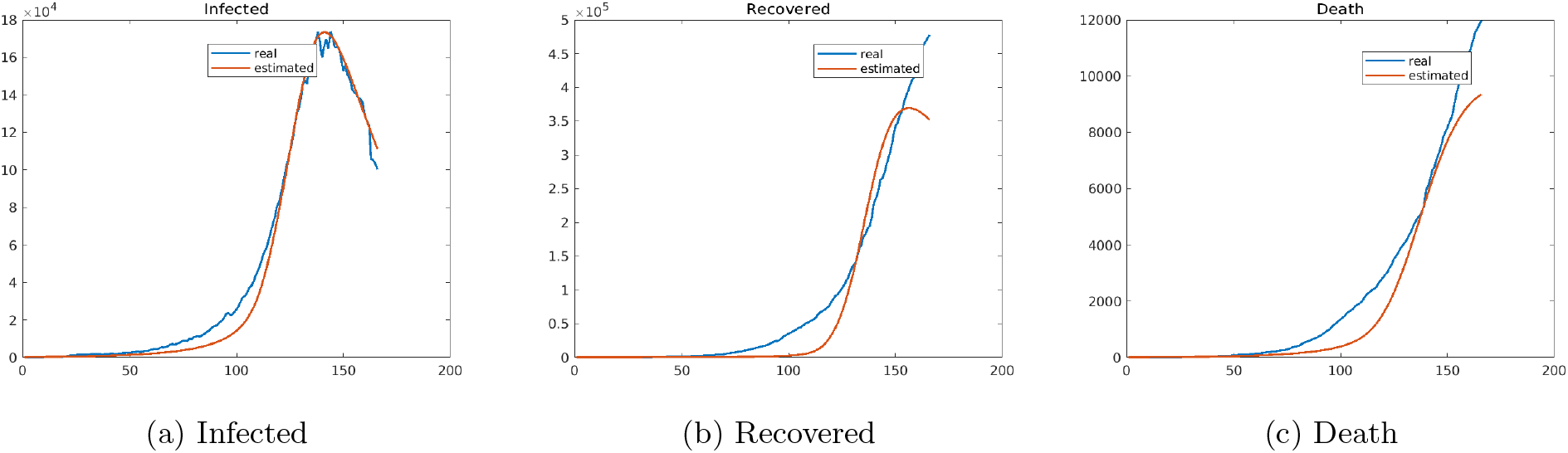
Numerical results of the fitted and observed values of Infected, Recovered and Death cases for South Africa.

The Figures in (4) shows the fitted model with the observed data for Brazil. We observe that the estimated parameters for the infected and recovered compartments are well fitted and approximated compared with the observed data. The death compartment has relatively small over predictions.

**Figure 4:**
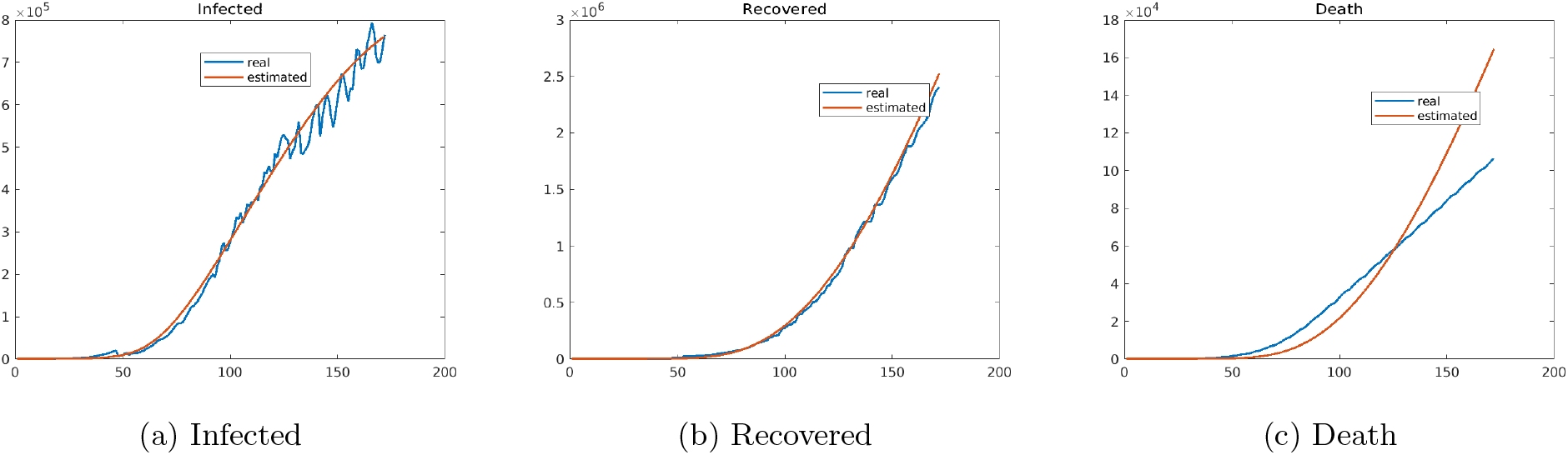
Numerical results of the fitted and observed values of Infected, Recovered and Death cases for Brazil.

The Figures (5) shows the fitted model with the observed values for Ethiopian data. We see that the estimated parameters for the infected and recovered compartments are well fitted and approximated compared with the observed data. It is also shown that the death compartment have relatively small over predictions in Ethiopia.

**Figure 5:**
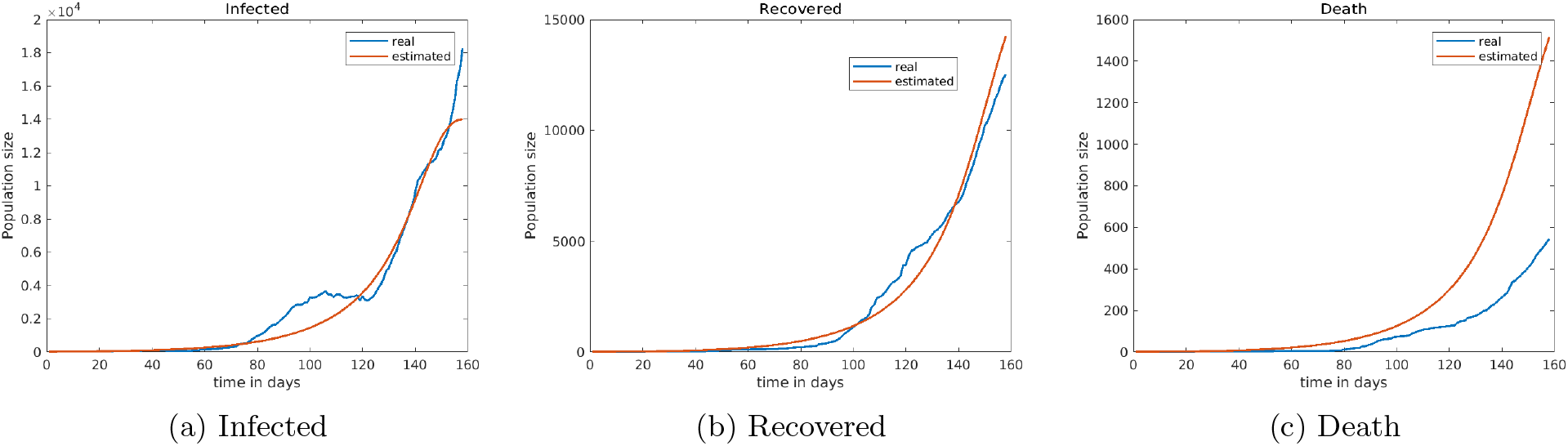
The numerical results of the fitted and observed values of Infected, Recovered and Death cases for Ethiopia.

### 4.3 Sensitivity of the Basic Reproduction Number

Sensitivity analysis is used to determine how sensitive a model is to changes in the value of the parameters and to changes in the structure of the model [3, 26]. In this paper, we focus on parameter sensitivity. Sensitivity analysis is a useful tool in model building as well as in model evaluation by showing how the model behavior responds to changes in parameter values [19].

#### Definition 4.1

*The Sensitivity and elasticity indices of the basic reproduction ratio, R*_0_ *, with respect to model parameter p are respectively given by* 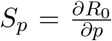 *and* 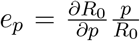*. That is, the elasticity indices is given by* 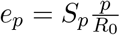 *[19, 17]*.

The graph in fig. 6 shows that in all countries the infection rates *β*_1_, *β*_2_, the recruitment rate *Λ*, the modification parameter *ν an*d shedding rate of the virus from the infected class to the environment *ξ ha*s the highest elasticity indices with COVID-19 being positively correlated to *R*_0_. An increase in these parameters will increase the spread of COVID-19 pandemic. The recovery rates *θ*, *γ*, the natural and virus-induced death rates *µ*, *δ*, the threshold value of the force of infection *λ*_0_, the pathogen concentration in the environment *K an*d the virus decay rate in the environment *ψ hav*e a negative correlation of *R*_0_ implying the virus decreases with an increase of these parameters.

**Figure 6:**
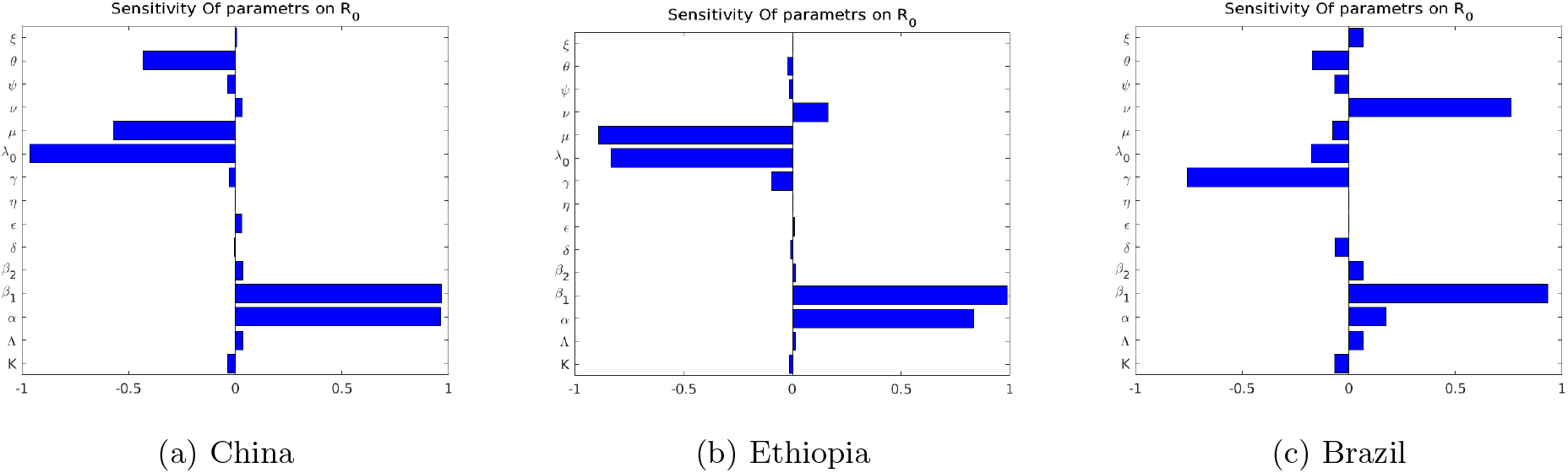
The visual representation of the elasticity indices of *R*_0_ with respect to the estimated parameters of countries China, Ethiopia and Brazil cases.

In Figure (7a), we observe that for the basic reproduction number *R*_0_ < 1, all solutions curve goes to the disease-free equilibrium point. These indicate that the disease-free equilibrium point is locally and globally asymptotically stable for the values of *R*_0_ < 1. In Figure (7b) with *R*_0_ < 1 and different initial conditions for the susceptible population, all trajectory goes to the equilibrium point 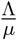 which indicates its stability.

**Figure 7:**
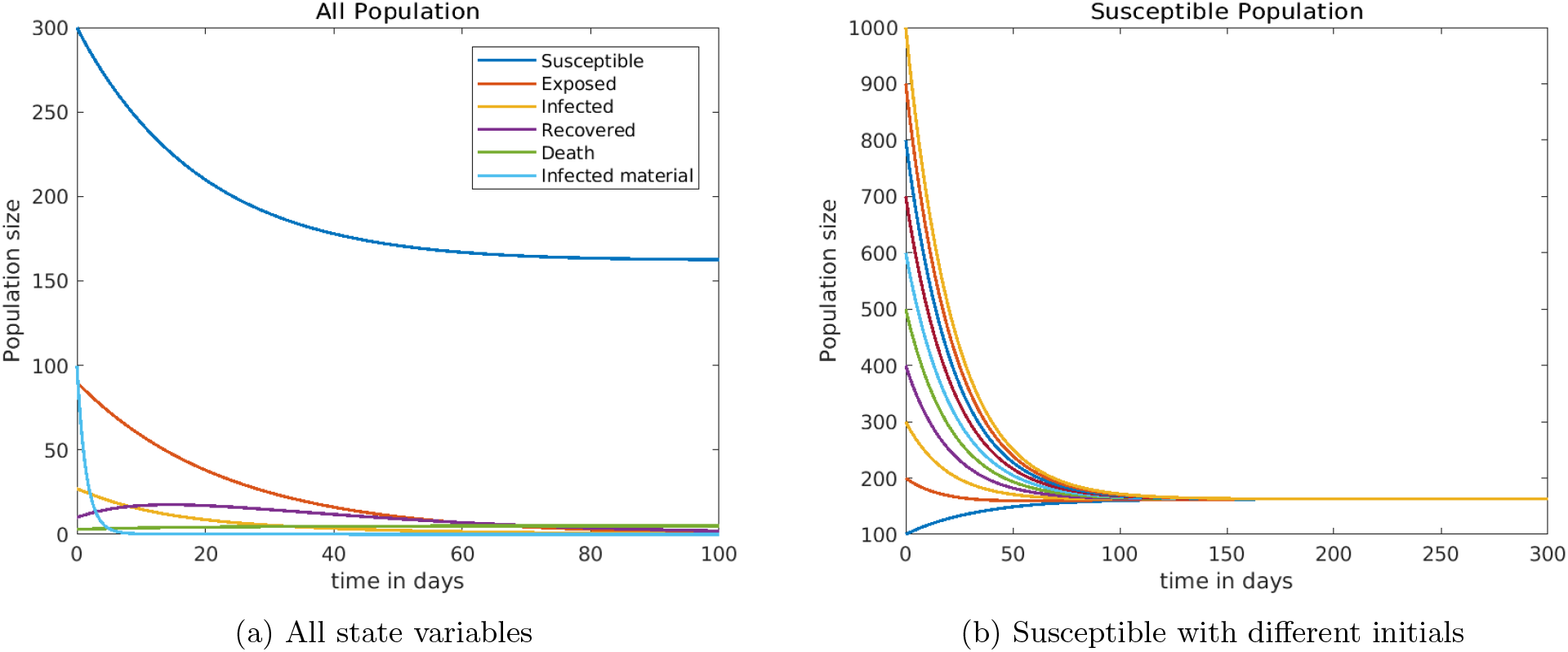
The trajectories of state variables for *R*_0_ =0.2552, which is less than one.

In Figure (8), we observe that for the basic reproduction number *R*_0_ *>* 1, all solutions curves goes away from the disease-free equilibrium point. These indicate that the disease-free equilibrium point is unstable for the values of *R*_0_ *>* 1, and the solutions will go to the endemic equilibrium point.

**Figure 8:**
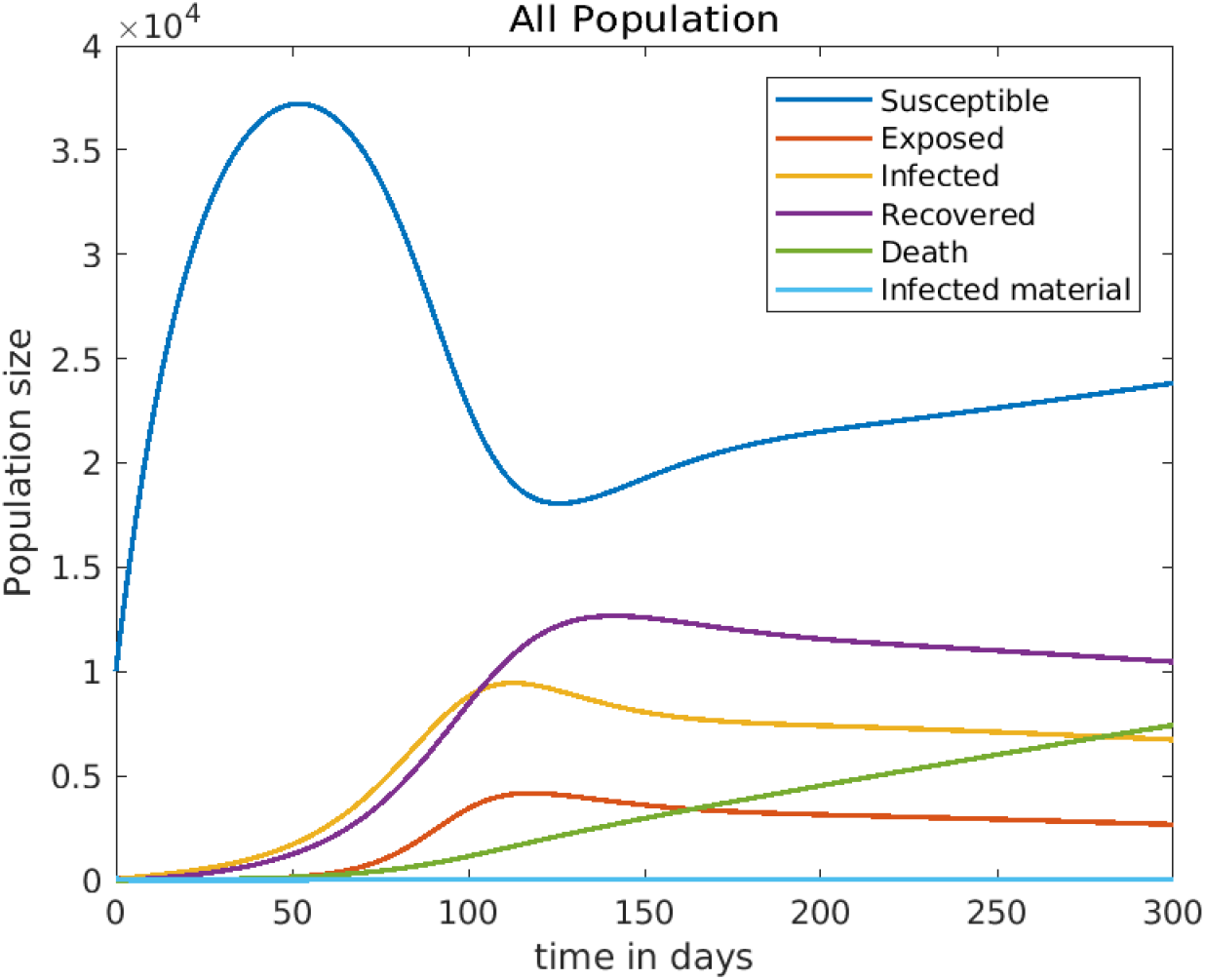
Trajectories of state variables for *R*_0_ =8.3636, which is greater than one.

### 4.4 Predictions

One of the most important applications of the dynamical system is to predict the future spread of the disease in the population. Here we predicted the number of active infections, recoveries and deaths for the countries Ethiopia, Brazil and South Africa for the next 45 days. In Figure 9, the fitted areas are similar to the Figures (3-5) while the predicted areas show the possible values of cases (active, recovered and death) up to September 30, 2020.

**Figure 9:**
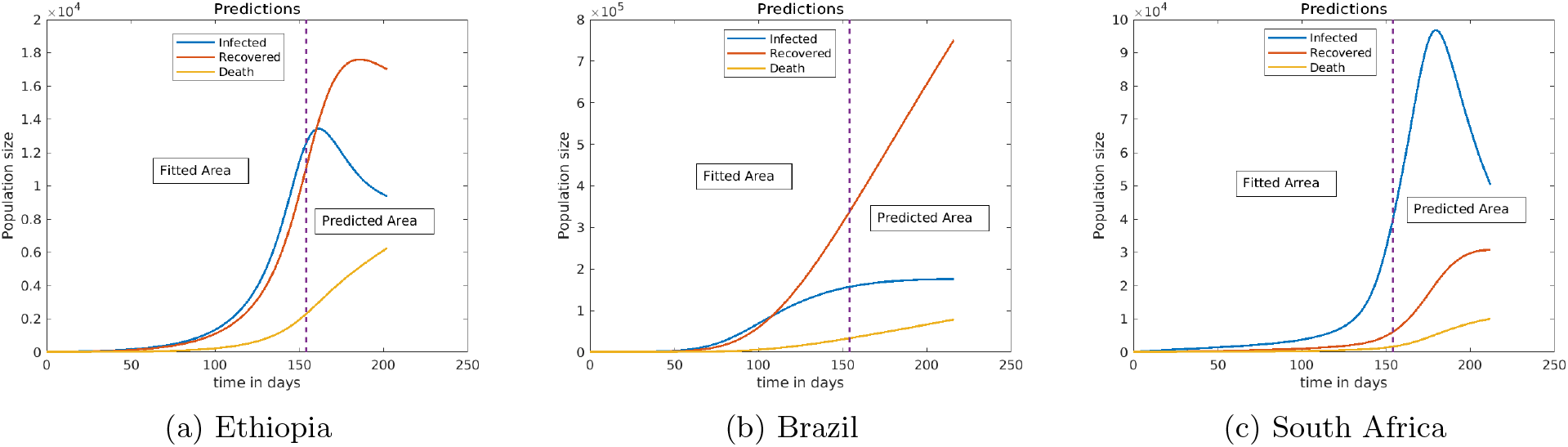
Predictions of active cases, recovered cases and Death Ethiopia, Brazil and South Africa on September 15, 2020.

## 5 Conclusion

In this paper, we have proposed and described a Susceptible-Exposed-Infected-Recovered-Death model with the addition of environmental infection with the virus for the transmission dynamics of the COVID-19 pandemic. We show the validity of the model by proving the existence, positivity and boundedness of the solutions. We then calibrated our mathematical model with a data-driven analysis, with the data coming from epidemiological results of WHO situation reports from 1 − 192. We programmed, simulated, and fitted the model with the observed data using Matlab. The estimation technique we use and the simulated results show a promising result.

For the disease-free equilibrium point, both the local and global stability analysis are proved and the result shows that the DFE is locally as well as globally asymptotically stable if *R*_0_ < 1 and unstable if *R*_0_ *>* 1.

The parameters are estimated using the combination of least square and Bayesian estimation techniques. According to our estimation, the model parameters vary from country to country as the case of the spread of virus varies accordingly. The sensitivity analysis on the countries show the infection rates *β*_1_ (human to human) and *β*_2_ (from the infected surfaces or environment) have high positive impacts for the spread of COVID-19. The threshold value of the force of infection for a population *λ*_0_, the recovery rates *θ*, *γ an*d the virus decay rate in the environment *ψ hav*e a negative impact on the spread of the virus. We have also observed that high numbers of people with knowledge about the virus, that are practicing the prescribed self-protective measures can slow down the outbreak.

It is recommended for individuals to increase their behaviors about the pandemic by following WHO recommendations such as using a face mask, practicing social distancing to decrease human to human transmission of the virus and washing their hands and infected surfaces with soap and alcohol-based sanitizer; which can decrease the transmission of the virus from the environment to humans and from infected individuals to the environment. It is also important to create awareness, disseminate information and change behavior of individuals to keep themselves which can reduce the pandemic threshold of the infections.

## Data Availability

The data in this study are available online on worldometer and WHO websites.

https://www.worldometers.info/coronavirus and https://www.who.int/emergencies/diseases/novel-coronavirus-2019/situation-reports

